# Discrepancies in patient and caregiver ratings of personality change in Alzheimer’s disease and related dementias

**DOI:** 10.1101/2023.03.09.23287003

**Authors:** Emma Rhodes, Dawn Mechanic-Hamilton, Jeffrey S. Phillips, Corey McMillan, Alejandra Bahena, Nykko Vitali, Quinn Hlava, Philip Cook, James Gee, Murray Grossman, Lauren Massimo

## Abstract

**Background:** Assessment of personality change in Alzheimer’s disease and related dementias (ADRD) is clinically meaningful but complicated by patient (i.e., reduced insight) and informant (i.e., caregiver burden) factors that confound accurate reporting of personality traits. This study assessed the impact of caregiver burden on informant report of Big Five personality traits (Extraversion, Agreeableness, Conscientiousness, Neuroticism, and Openness) and investigated regional cortical volumes associated with larger discrepancies in patient and informant report of Big Five personality traits.

**Methods:** Sixty-four ADRD participants with heterogeneous neurodegenerative clinical phenotypes and their informants completed the Big Five Inventory (BFI). Caregiver burden was measured using the Zarit Burden Interview (ZBI). Discrepancy scores were computed as the absolute value of the difference between patient and informant ratings for all BFI trait scores and summed to create a global score. Regional grey matter volumes from T1-weighted 3T MRI were normalized to intracranial volume and related to global Big Five discrepancy scores using linear regression.

**Results:** Higher levels of caregiver burden were associated with higher informant ratings of patient Neuroticism (ß =0.27, p =.016) and lower informant ratings of patient Agreeableness (ß =-0.32, p =.002), Conscientiousness (ß =-0.3, p =.002), and Openness (ß =-0.34, p =.003) independent of disease severity. Patients with greater Big Five discrepancy scores showed smaller cortical volumes in right medial PFC (β = -0.00015, *p* = .002), right superior temporal gyrus (β = -0.00028, *p* = .025), and left inferior frontal gyrus (β = -0.00006 *p* = .013).

**Conclusions:** Informant ratings of personality traits in ADRD can be confounded by caregiver burden, highlighting the need for more objective measures of personality and behavior in dementia samples. Discrepancies between informant and patient ratings of personality may additionally reflect loss of insight secondary to cortical atrophy in frontal and temporal structures.

---

Personality change is a common feature of Alzheimer’s disease and related dementias (ADRD) with distinct patterns and severity across neurodegenerative phenotypes and evidence to suggest that degree of personality change reflects underlying disease progression (Helmes et al., 2013; Rankin et al., 2003; Sollberger et al., 2011). However, measurement of personality in ADRD is compromised by patient and caregiver factors that can confound accurate reporting of personality traits, such as reduced insight and cognitive impairment in patients and elevated burden and distress in caregivers (Lechner & Rammstedt, 2015). Discrepancy scores measuring the difference between patient and informant report of personality in individuals with ADRD can provide valuable information on patient? insight into personality change (Rankin et al., 2005), but previous research has failed to account for caregiver factors such as elevated burden that can confound informant report of personality change. Caregiver burden has been shown to bias informant report of personality in patients with Alzheimer’s disease (AD; Welleford et al., 1995), but the extent to which burden impacts caregiver report of personality traits in other dementia phenotypes remains unclear. This study was conducted to assess patient and caregiver factors that contribute to discrepancies in report of Big Five personality traits (Extraversion, Agreeableness, Conscientiousness, Neuroticism, Openness) in a mixed ADRD sample.

The Big Five model of personality defines five global traits of human personality: 1) Extraversion, or a tendency to be outgoing and engaged with the external world; 2) Agreeableness, or a tendency to be compassionate, generous, and trustworthy with others; 3) Conscientiousness, or the tendency to be disciplined and strive for achievement; 4) Neuroticism, or a tendency to be vulnerable to stress and experience negative emotions (e.g., sadness, anxiety); and 5) Openness to new experiences, or a tendency to be curious, open to emotion, and willing to try new things (see McCrae & Costa Jr., 1999 for review). Studies examining change in Big Five personality traits in ADRD have shown an increase in Neuroticism and decrease in Extraversion, Openness and Conscientiousness in individuals with AD (Siegler et al., 1994), as well as increased Neuroticism and decreased Conscientiousness in mild cognitive impairment (MCI) and very mild AD (Duchek et al., 2007). Findings from frontotemporal dementia (FTD) consistently show reduced Conscientiousness relative to healthy controls and AD comparison samples, while there is mixed evidence of both increased and decreased Neuroticism (Lykou et al., 2013; Mahoney et al., 2011). However, these findings are limited by reliance on informant report of personality, which can be confounded by elevated burden in caregivers for individuals with AD. The extant literature is sparse, but one study using a sample of AD caregivers found that higher levels of burden were associated with lower informant report of conscientiousness and higher report of neuroticism, independent of disease severity (Welleford et al., 1995). Thus, it is not clear to what degree informant reported changes in Big Five traits reflect elevated caregiver burden versus objective personality change, and less is known about the impact of caregiver burden on informant report of personality in other ADRD phenotypes.

Neuroimaging research has highlighted a biological basis for personality traits in healthy adults and persons with ADRD. Structural MRI studies in healthy older adults have linked Big Five personality traits with a distributed network of frontal and temporal cortical volumes including the dorsolateral prefrontal cortex (DLPFC; associated with Neuroticism and Extraversion), medial orbitofrontal cortex (mOFC; associated with all 5 personality traits), anterior cingulate cortex (ACC; associated with Extraversion), medial temporal lobe (associated with Conscientiousness), frontal operculum (associated with Neuroticism), and insula (associated with Openness; DeYoung et al., 2010; Jackson et al., 2011; Kapogiannis et al., 2013). In ADRD, personality change has been linked to cortical atrophy in predominantly right-sided frontal and temporal regions, including the superior temporal gyrus, DLPFC, mOFC, and ACC (Mahoney et al., 2011; Sollberger et al., 2009), many of which have also been implicated in anosognosia and reduced self-awareness in ADRD. Specifically, evidence from structural neuroimaging has identified atrophy in the right ventromedial prefrontal (vmPFC), frontopolar, OFC, ACC, insula, amygdala, and retrosplenial cortex as associated with reduced insight in FTD and AD (Hornberger et al., 2012; Rosen et al., 2010), but it remains unclear if the neural substrates of reduced insight into personality reflect more than a general pattern of anosognosia in ADRD. Further, while discrepancy scores have been used widely to capture loss of insight into various domains of functioning (e.g., memory, social cognition, disability) in ADRD, no existing studies have accounted for the confounding effect of caregiver burden when examining neuroanatomic underpinnings of reduced insight.

This study had two aims. The first aim was to evaluate discrepancies in patient and informant report of Big Five personality traits in a mixed dementia sample. We hypothesized that patients would rate themselves higher on positive traits (Extraversion, Agreeableness, Conscientiousness, Openness) and lower on Neuroticism relative to their informants. The second aim was to assess sources of discrepancy between patient and informant report of Big Five personality traits. We hypothesized that caregiver burden would significantly contribute to discrepancies in reporting and that larger discrepancies would be associated with cortical volumes in right-sided frontal (OFC, ACC, mPFC) and temporal (Amygdala, STG) brain regions implicated in insight and self-awareness.

## Methods

### Participants

The sample included 64 participants diagnosed with a heterogeneous mix of ADRD clinical diagnoses (Table 1) who were recruited from the University of Pennsylvania Frontotemporal Degeneration Center (UPenn FTDC). We included a mix of ADRD phenotypes to ensure a broad range of insight and to assess the neuroanatomic specificity of regional brain volumes associated with insight compared to the extant literature, which has largely focused on insight within individual ADRD phenotypes. All patients underwent a comprehensive clinical evaluation including a clinical history obtained from patients and caregivers, neurologic examination, mental status exam, neuropsychological assessment, and 3T structural MRI. Clinical diagnosis was made by consensus of at least two trained reviewers using published criteria for AD (McKhann et al., 2011), bvFTD (Rascovsky et al., 2011), nonfluent/agrammatic primary progressive aphasia (naPPA), semantic variant PPA (svPPA), logopenic variant PPA (lvPPA; Gorno-Tempini et al., 2011), Parkinson’s disease with dementia (PDD), dementia with Lewy bodies (DLB; Litvan et al., 2003), corticobasal syndrome (CBS; Armstrong, et al., 2013), progressive supranuclear palsy (PSP; Hoglinger, et al., 2017), and mild cognitive impairment (MCI; Petersen et al., 2014). All participants completed an informed consent procedure approved by the University of Pennsylvania Institutional Review Board as well as personality assessment using the Big Five Inventory. Inclusion criteria included a consensus diagnosis of neurodegenerative disease, mild-moderate disease severity defined by Mini Mental State Examination (MMSE; Folstein et al., 1975) total score > 15, and the presence of a cognitively unimpaired study partner to provide informant report of clinical and personality factors. Exclusion criteria included the presence of other neurologic conditions such as stroke, traumatic brain injury, or hydrocephalus (determined by clinical history and neuroimaging studies) or psychiatric disorders such as major depressive disorder or schizophrenia. A separate group of age-matched healthy controls (HC) was used to establish a pattern of regional cortical atrophy in the ADRD sample. HCs were recruited from the UPenn FTDC, and inclusion criteria included unimpaired performance on the Telephone Interview for Cognitive Status (TICS Total Score > 130). Exclusion criteria included a history of dementia, mild cognitive impairment, or any other neurologic or psychiatric condition that could affect cognitive and behavioral functioning.

**Table 1.**
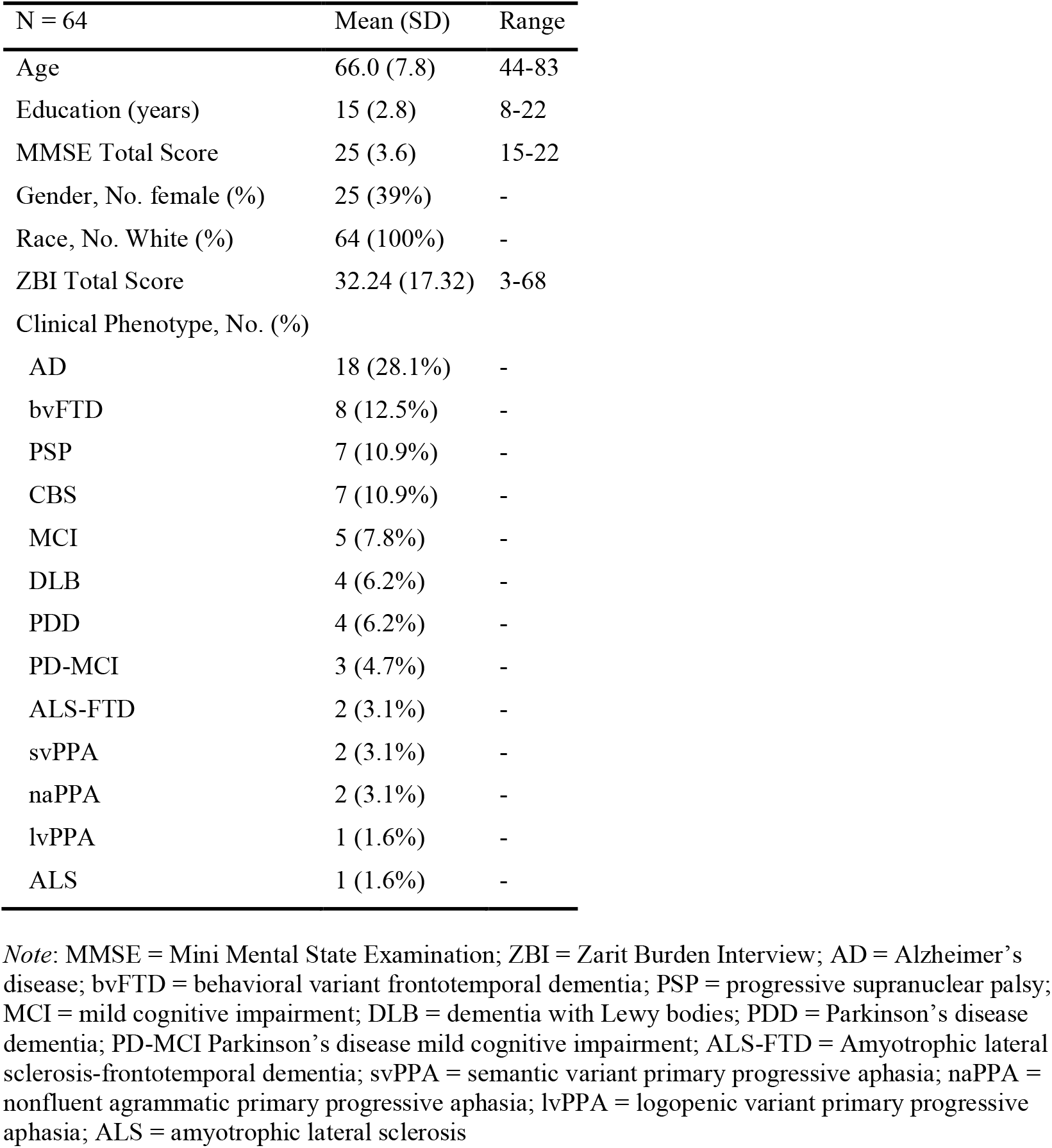
Patient Sample Characteristics

### Measures

#### Personality

Patients and informants completed the 44-item Big Five Inventory (BFI; John et al., 1991), which assesses personality domains outlined in the Big Five Model of personality (Extraversion, Agreeableness, Conscientiousness, Neuroticism, and Openness). BFI items consist of short phrases that are rated by on a 5-point Likert scale ranging from “Disagree strongly” to “Agree strongly”, with eight to ten items per scale. Scores for each factor are computed as the mean rating across relevant items, with a minimum score of 1 and a maximum score of 5. The BFI has demonstrated strong structural, convergent, and divergent validity (John et al., 1991; Benet-Martínez & John, 1998) and has been used to identify change in personality in ADRD (Mahoney et al., 2011; Mychack et al., 2001). Discrepancy scores were calculated for each participant-informant dyad as the absolute value of the difference between patient self-ratings and informant ratings of patient personality across all Big Five personality factors. A global discrepancy score representing the overall disagreement between patient and caregiver on personality ratings was computed as the sum of the five individual discrepancy scores.

#### Disease Severity

Disease severity was approximated using the MMSE, a measure of global cognitive functioning (Folstein et al., 1975) administered by trained research personnel. Possible scores range from 0 to 30 with higher scores reflecting better global cognitive performance.

#### Caregiver Burden

Caregiver burden was measured using the Zarit Burden Interview (ZBI, Zarit & Zarit, 1987), which measures perceived burden of caregivers with 22 items on a 5-point Likert scale. Scores on all items were summed to produce a total score ranging from 0 to 88 with higher scores reflecting greater caregiver burden. The ZBI has demonstrated good reliability and validity in dementia caregivers (Seng et al., 2010).

### Procedures

#### Structural Neuroimaging

T_1_-weighted magnetic resonance images were acquired for 65 patients and 26 age-matched controls using a Siemens 3-Tesla MRI scanner. Eleven scans were acquired sagittally with voxel sizes ranging from 0.98 mm × 0.98 mm × 1 mm to 1.25 mm × 1.25mm × 1 mm; repetition times of 2300 ms to 3000 ms; and echo times of 0.00254 ms to 0.00355 ms. The remaining 80 images were acquired axially with 0.98 mm × 0.98 mm × 1 mm voxels, a 256 × 192 matrix, a repetition time of 1620 ms, an echo time of 0.00309 ms, and a flip angle of 15°. Scans were visually inspected for quality by three raters. Disagreements between raters were resolved by a consensus amongst all three raters. Advanced Normalization Tools (ANTs) was used to process each image as previously described (Avants et al., 2014; Tustison et al., 2014). Briefly, all images were stripped of the skull and underwent intensity normalization (Tustison et al., 2010). Images were segmented into six tissue classes (cortical grey matter, deep grey matter, white matter, CSF, brainstem, and cerebellum) using template-based priors; this tissue segmentation was then used to estimate volumes. We used the 100 parcels, 17 network atlas of Schaefer and colleagues (Schaefer et al., 2018) to calculate the volume of grey matter voxels within each label in each subject’s native T1 space. Label volumes were normalized by intracranial volume prior to use in regression analyses, which was estimated from brain mask volume. Normalized cortical volumes for the healthy control group were used as a reference group to identify regions of relative cortical atrophy in the overall patient group.

#### Statistical Analyses

Continuous variables were summarized with mean and SD. Categorical variables were summarized with percentages. Group differences in caregiver and patient reported personality traits were assessed using one-way ANOVA. Multiple linear regressions were used to assess the independent association of caregiver burden on caregiver Big Five personality ratings covarying for patient age, education, MMSE. We additionally covaried for patient Big Five self-ratings in order to account for shared variance between patient and informant ratings that may reflect accurate and unbiased report of personality. Independent samples t-tests were used to identify regions of cortical atrophy in the patient group relative to age-matched healthy controls using Holm adjustment for multiple comparisons (Supplemental Table 1). The Big Five global discrepancy score was then regressed onto regions showing smaller cortical volumes in patients relative to healthy controls covarying for age, sex, education, ZBI total score, and MMSE total score with Holm adjustment. Additional models were performed examining the impact of total individual discrepancy scores for each Big Five traits, but results did not survive correction for multiple comparisons.

## Results

### Sample Characteristics

The patient sample was clinically and demographically heterogeneous except for racial and ethnic background, which was 100% white and non-Hispanic (Table 1). Patients met diagnostic criteria for a variety of neurodegenerative clinical phenotypes, with AD and FTLD (bvFTD, naPPA, svPPA, ALS-FTD) accounting for half of the sample (32/64). Other clinical diagnoses included in the sample were PDD, DLB, CBS, PSP, MCI, and PD-MCI. Mean age at testing was 65.84 (SD = 7.8), 39% of the sample were women (n = 25), and mean MMSE was 25.67 (SD = 3.57). Seventy percent (n = 45) of caregivers were female; 89.2% (n = 58) were spouses or partners, 6.15% (n = 4) were adult children or stepchildren, and one caregiver was a friend. Caregiver demographic information was missing for one participant. Mean caregiver ZBI was 31.29 (SD = 17.37), which is in the moderate range (Zarit & Zarit, 1987).

### Patient and Caregiver Report of Big Five Personality Traits

Caregivers rated patients lower on Extraversion (F[63, 1] = 7.15, *p* = .008), Conscientiousness (F[63, 1] = 26.33, p < .001), and Openness (F[63, 1] = 14.23, *p* < .001) and higher on Neuroticism (F[63, 1] = 12.75, *p* < .001) relative to patients’ self-ratings. There was no difference between patient and caregiver ratings of Agreeableness (*p* > .05; Table 2).

**Table 2.** Patient and Caregiver Report of Big Five Personality Factors

|  | Patient | Caregiver | Discrepancy<br>(Patient –<br>Caregiver) |  |  |
| --- | --- | --- | --- | --- | --- |
|  | Mean (SD) | Mean (SD) | Mean (SD) | F | *p*-value |
| Extraversion | 3.34 (0.77) | 2.91 (1.0) | 0.43 (0.93) | 7.153 | **0.008** |
| Agreeableness | 4.2 (0.6) | 4.08 (0.82) | 0.18 (0.88) | 0.858 | 0.356 |
| Conscientiousness | 3.94 (0.61) | 3.21 (0.95) | 0.64 (0.99) | 26.33 | **< .001** |
| Neuroticism | 2.48 (0.83) | 3.01 (0.85) | -0.48 (0.92) | 12.75 | **< .001** |
| Openness | 3.43 (0.61) | 2.89 (0.97) | 0.58 (0.91) | 14.23 | **< .001** |

Note: SD = standard deviation

After covarying for patient age, education, MMSE total score, and patient Big Five ratings, higher ZBI total score was significantly associated with lower caregiver ratings of patient Agreeableness (β = -0.32, *p* = .002), Conscientiousness (β = -0.30, *p* = .002), and Openness (β = -0.34, *p* = .003) as well as higher ratings of patient Neuroticism (β = 0.27, *p* = .016; Table 3). ZBI total score was not associated with caregiver ratings of Extraversion (*p* > .05).

**Table 3.** Associations of Age, Education, Global Cognition, Caregiver Burden, and Patient Big Five Ratings with Caregiver Big Five Ratings

|  | Extraversion | Agreeableness | Conscientiousness | Neuroticism | Openness |
| --- | --- | --- | --- | --- | --- |
| | $\beta$ (SE) | $\beta$ (SE) | $\beta$ (SE) | $\beta$ (SE) | $\beta$ (SE) |
| (Intercept) | **2.93** (0.13)*** | **4.08** (0.11)*** | **3.17** (0.12)*** | **3.07** (0.11)*** | **2.84** (0.11)*** |
| Age | 0.24 (0.15) | -0.02 (0.12) | 0.11 (0.14) | 0.13 (0.12) | 0.02 (0.13) |
| Education | 0.15 (0.15) | 0.01 (0.12) | 0.15 (0.14) | 0.03 (0.13) | 0.17 (0.13) |
| MMSE | 0.08 (0.14) | 0.00 (0.12) | -0.04 (0.13) | -0.01 (0.11) | 0.08 (0.12) |
| ZBI | 0.06 (0.15) | **-0.32 \*** (0.12) | **-0.3 \*** (0.14) | **0.27 \*** (0.12) | **-0.34 \*\*** (0.13) |
| Patient Rating | **0.44 \*\*** (0.14) | **0.25 \*** (0.12) | **0.36 \*\*** (0.13) | 0.18 (0.12) | **0.39 \*\*** (0.13) |
| R2 | 0.23 | 0.24 | 0.28 | 0.15 | 0.45 |

Note: \*\*\* $p < 0.001$ ; \*\* $p < 0.01$ ; \* $p < 0.05$ ; SE = standard error; MMSE = Mini Mental State Examination; ZBI = Zarit Burden Interview

### Big Five Discrepancy Scores and Regional Cortical Volumes

Relative to an age-matched sample of healthy controls, the overall ADRD sample showed smaller cortical volumes in the bilateral somatomotor cortex, left frontal eye fields, left postcentral gyrus, left inferior frontal gyrus, right medial and ventromedial PFC, right superior temporal gyrus, and left superior parietal lobule (Supplemental Table 1). These regions were treated as dependent variables in separate multiple regression models assessing associations of the global Big Five discrepancy score with lateralized regional cortical volumes covarying for age, sex, education, and MMSE total score (Tables 4 & 5). After correction for multiple comparisons, we observed that greater global Big Five discrepancy scores were associated with smaller cortical volumes in the right medial PFC (β = -0.00015, *p* = .002), right superior temporal gyrus (β = -0.00028, *p* = .025), and left inferior frontal gyrus (β = -0.00006 *p* = .013; Figure 1). No other ROIs showed a significant association to the global Big Five discrepancy score (all *p*’s > .05).

**Table 4.** Associations of right hemisphere cortical volumes with global Big Five discrepancy scores

|  | Right Superior Parietal |  |  | Right Somatomotor |  |  | Right Ventromedial PFC |  |  | Right Medial PFC |  |  | Right Superior Temporal |  |  |
| --- | --- | --- | --- | --- | --- | --- | --- | --- | --- | --- | --- | --- | --- | --- | --- |
|  | R<sup>2</sup> | DF | p | R<sup>2</sup> | DF | p | R<sup>2</sup> | DF | p | R<sup>2</sup> | DF | p | R<sup>2</sup> | DF | p |
|  | 0.07 | 5, 52 | 0.13 | 0.14 | 5, 52 | 0.218 | 0.05 | 5, 52 | 0.179 | 0.19 | 5, 52 | 0.003 | 0.13 | 5, 52 | 0.011 |
|  | β | SE | p | β | SE | p | β | SE | p | β | SE | p | β | SE | p |
| Intercept | 0.0051 | 0.00223 | 0.02 | 0.00165 | 0.00048 | 0.001 | 0.00389 | 0.00113 | 0.001 | 0.00561 | 0.00127 | < 0.001 | 0.00389 | 0.00113 | 0.001 |
| Age | -0.00001 | 0.00002 | 0.99 | -0.00001 | 0.0 | 0.029 | -0.00001 | 0.00001 | 0.48 | -0.00001 | 0.00001 | 0.49 | -0.00001 | 0.00001 | 0.48 |
| Education | -0.00007 | 0.00005 | 0.30 | -0.00002 | 0.00001 | 0.19 | -0.00002 | 0.00003 | 0.52 | -0.00002 | 0.00003 | 0.52 | -0.00002 | 0.00003 | 0.52 |
| MMSE | 0.00002 | 0.00004 | 0.40 | 0.00001 | 0.00001 | 0.27 | 0.0 | 0.00002 | 0.96 | 0.00002 | 0.00002 | 0.28 | 0.0 | 0.00002 | 0.96 |
| ZBI Total | 0.0 | 0.00001 | 0.89 | 0.0 | 0.0 | 0.73 | -0.00001 | 0.0 | 0.12 | -0.00002 | 0.00001 | 0.008 | -0.00001 | 0.0 | 0.12 |
| Global Big 5 Discrepancy | 0.00012 | 0.00007 | 0.06 | 0.00001 | 0.00001 | 0.65 | -0.00006 | 0.00004 | 0.11 | **-0.00015** | **0.00003** | **0.002** | **-0.00028** | **0.00004** | **0.025** |

**Table 5.** Associations of left hemisphere cortical volumes with global Big Five discrepancy scores

|  | Left Somatomotor |  |  | Left Postcentral |  |  | Left Frontal Eye Fields |  |  | Left Inferior Frontal |  |  |
| --- | --- | --- | --- | --- | --- | --- | --- | --- | --- | --- | --- | --- |
|  | R<sup>2</sup> | DF | p | R<sup>2</sup> | DF | p | R<sup>2</sup> | DF | p | R<sup>2</sup> | DF | p |
|  | 0.12 | 5, 52 | 0.13 | 0.02 | 5, 52 | 0.513 | 0.081 | 5, 52 | 0.94 | 0.16 | 5, 52 | 0.004 |
|  | β | SE | p | β | SE | p | β | SE | p | β | SE | p |
| Intercept | 0.0026 | 0.00163 | 0.08 | 0.0017 | 0.00085 | 0.50 | 0.0031 | 0.00197 | 0.07 | 0.0014 | 0.00067 | 0.021 |
| Age | 0.00002 | 0.00004 | 0.61 | -0.00001 | 0.00001 | 0.44 | 0 | 0.00002 | 0.98 | 0.00001 | 0 | 0.17 |
| Education | 0.00001 | 0.00002 | 0.59 | -0.00003 | 0.00002 | 0.28 | 0 | 0.00005 | 0.93 | -0.00001 | 0.00002 | 0.47 |
| MMSE | 0 | 0.00001 | 0.85 | 0.00001 | 0.00002 | 0.44 | 0.00003 | 0.00003 | 0.36 | 0.00002 | 0.00001 | 0.11 |
| ZBI Total | 0 | 0.00001 | 0.95 | 0 | 0 | 0.5 | 0 | 0.00001 | 0.76 | -0.00001 | 0 | 0.029 |
| Global Big 5<br>Discrepancy | -0.00001 | 0.00002 | 0.64 | 0.00003 | 0.00002 | 0.19 | -0.00001 | 0.00004 | 0.88 | **-0.00006** | **0.00002** | **0.013** |

**Figure 1.**
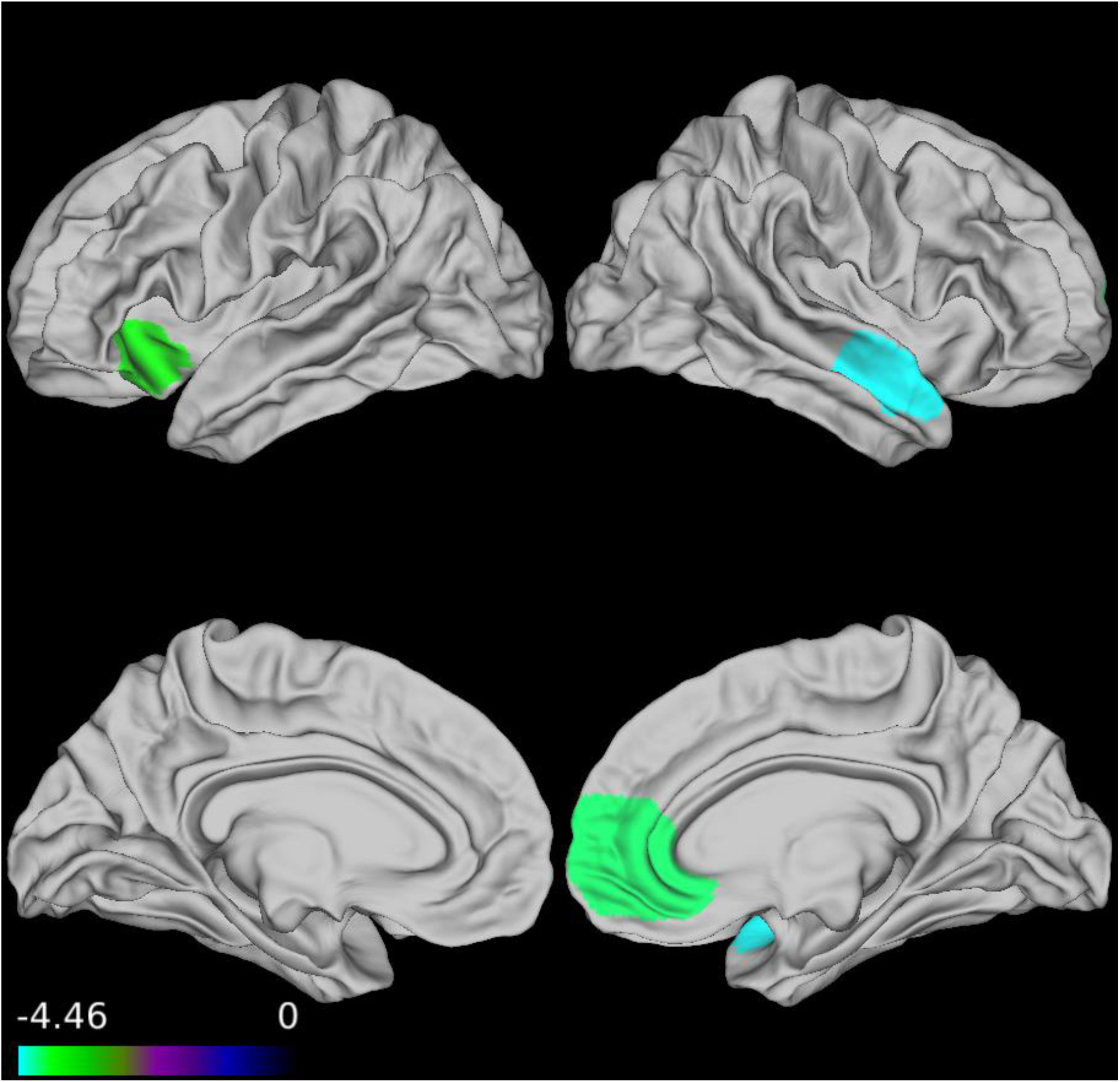
Heat map of normalized cortical volume ROIs associated with Big Five Global discrepancy score

## Discussion

This project assessed and investigated sources of discrepancies in patient and informant report of Big Five personality traits in ADRD, namely caregiver burden and patient insight. We found that ADRD patients reported lower levels of Neuroticism and higher levels of Extraversion, Conscientiousness, and Openness relative to their caregivers, and that caregiver burden influenced informant ratings of Conscientiousness, Neuroticism, Openness, and Agreeableness. We also found that patients with greater discrepancies between self and informant ratings of personality showed smaller cortical volume in the right mPFC, right superior temporal gyrus, and left inferior frontal gyrus, areas that underlie insight and self-awareness in ADRD and healthy adults. Each of these findings will be discussed in detail below.

Consistent with our hypotheses and findings from previous work (Sollberger et al., 2014; Rosen, et al., 20010; Hornberger et al., 2012) using other personality rating scales, we found that ADRD patients showed a pattern of self-report in which a negative trait (e.g., Neuroticism) was lower and positive traits (e.g., Conscientiousness, Openness, Extraversion) were higher relative to caregiver report. We expected that discrepancies in patient and caregiver report of personality may be due to multiple factors, including loss of insight into personality change and bias in caregiver report due to high levels of burden. When we investigated the independent impact of caregiver burden on informant report of patent personality, we found that higher burden was associated with lower informant ratings of positive traits (Agreeableness, Conscientiousness, Openness) and higher informant ratings of Neuroticism, suggesting that caregiver ratings of unfavorable personality change may, in part, reflect distress secondary to high levels of burden. This is consistent with previous work in AD showing that overburdened caregivers are more likely to perceive and report more negative personality change in care receivers (Welleford et al., 1995).

Alternatively, caregiver burden may be higher in informants of patients with greater reductions in Agreeableness, Conscientiousness, and Openness and a greater increase in Neuroticism, as these patients may be more difficult to care for. Of note, our informant sample reported a moderate level of burden on average, which is consistent with previously reported levels of burden in mixed dementia samples (Springate & Tremont, 2014) and suggests a severe level of burden is not necessary to impact ratings of personality change.

Reduced insight into personality change can also contribute to greater discrepancy scores, as patients with limited insight may rate themselves more favorably or more like their premorbid personality compared to informant ratings (Lechner & Rammstedt, 2015). We observed that greater discrepancies in overall Big Five ratings between patient and caregiver were associated with smaller cortical volumes in frontal (right mPFC, left inferior frontal gyrus) and temporal (right superior temporal gyrus) regions that have been implicated in insight and self-awareness in ADRD. Our right-lateralized findings are consistent with prior work showing right-sided frontal and temporal lobe contributions to personality and insight in healthy adults and individuals with ADRD (Kapogiannis et al., 2013; Mychack et al., 2001). Specifically, the right mPFC, including the ventral portion of the ACC, has been linked to personality change and poor self-awareness in bvFTD (Massimo et al., 2013; Rosen et al., 2005), and the right superior temporal gyrus has been linked to awareness of behavioral disturbances, self-perception, and self-representation across various ADRD phenotypes (Ruby et al., 2007; Sollberger et al., 2014; Zamboni et al., 2010). Further, we found that patients with larger discrepancies in personality reporting had smaller volumes in the left inferior frontal gyrus, which is most commonly associated with speech and language production.

However, evidence from healthy adults suggests that this region is also implicated in self-awareness via production of so called “inner speech,” which is thought to be a prerequisite for introspective and self-referential processes (Morin, 2005; Morin & Michaud, 2007). Taken together, our imaging results provide additional support for the right mPFC and superior temporal gyrus as key regions implicated in insight in ADRD and highlight a novel role for the left inferior frontal gyrus in self-awareness of personality change that may reflect the impact of self-referential language.

This is the first study to our knowledge to investigate discrepant perceptions of Big Five personality traits in persons with ADRD and their caregivers. Though our findings are bolstered by the use of a well-characterized sample, the heterogeneity of clinical phenotypes with unequal representation across groups precluded subgroup analyses that may have been informative in assessing the uniformity of these findings across phenotypes. Additionally, the generalizability of our work is limited by a racially homogenous and highly educated sample, and we were limited in our measurement of disease severity and relied on the MMSE, which is a gross estimation of functioning and can be confounded by phenotypically specific cognitive deficits (e.g., lower scores due to language impairment in PPA subtypes). Further, our data did not include specific measurement of insight or self-awareness, which would have strengthened our conclusions about the relative contribution of patient insight on Big Five discrepancy scores. Future work should explore the contributions of specific cognitive impairments (e.g., comprehension of test items) and abilities (e.g., inner speech) on patient report of personality and other domains of functioning (e.g., ADLs, mood).

This study was conducted to assess the impact of patient and caregiver factors on the report of personality traits in a mixed ADRD sample. We found that caregiver burden contributed to less favorable informant ratings of patient personality, and that patients whose self-rating differed most from their informant’s rating showed evidence of greater atrophy in the right mPFC and superior temporal gyrus and left inferior frontal gyrus. Together, our findings contribute to a growing literature highlighting the confounding effects of individual patient and caregiver characteristics on informant report of personality and other behavioral features of dementia and underscores the need for alternative and more objective methods of measuring non-cognitive symptoms of ADRD.

## Data Availability

All data produced in the present study are available upon reasonable request to the authors

