## Supplemental Table 1 for "Discrepancies in patient and caregiver ratings of personality change in Alzheimer’s disease and related dementias"

| **Table 4.** Effect of Big 5 Global Discrepancy on Right Hemisphere Cortical Volumes | | | | | | | | | | | | | | | | | | | | | | | | | | | | | |
| --- | --- | --- | --- | --- | --- | --- | --- | --- | --- | --- | --- | --- | --- | --- | --- | --- | --- | --- | --- | --- | --- | --- | --- | --- | --- | --- | --- | --- | --- |
|  | **Right Superior Parietal** | | | |  | **Right Somatomotor** | | | |  | **Right Ventromedial PFC** | | | |  | **Right Medial PFC** | | |  | | |  | **Right Superior Temporal** | | | | | | |
|  | R2 | 0.073 |  |  | | R2 | 0.144 |  |  | | R2 | 0.056 |  |  | | R2 | 0.322 |  | | |  | | R2 | 0.131 | |  | |  | |
|  | F | 1.795 |  |  | | F | 1.46 |  |  | | F | 1.6 |  |  | | F | 5.76 |  | | |  | | F | 1.11 | |  | |  | |
|  | DF | 5, 52 |  |  | | DF | 5, 52 |  |  | | DF | 5, 52 |  |  | | DF | 5, 52 |  | | |  | | DF | 5, 52 | |  | |  | |
|  | p | 0.133 |  |  | | p | 0.218 |  |  | | p | 0.179 |  |  | | p | **0.00036** |  | | |  | | p | **0.0113** | |  | |  | |
|  | β | SE | t | p | | β | SE | t | p | | β | SE | t | p | | β | SE | t | | p | | | β | | SE | | t | | p |
| Intercept | 0.00508 | 0.00218 | 2.33 | 0.02 | | 0.00165 | 0.00048 | 3.41 | 0.001 | | 0.00389 | 0.00113 | 3.46 | 0.001 | | 0.00561 | 0.00127 | 4.42 | | < 0.001 | | | 0.00389 | | 0.00113 | | 3.46 | | 0.001 |
| Age | 0 | 0.00002 | 0.00 | 0.99 | | -0.00001 | 0 | -2.26 | 0.029 | | -0.00001 | 0.00001 | -0.72 | 0.48 | | -0.00001 | 0.00001 | -0.70 | | 0.49 | | | -0.00001 | | 0.00001 | | -0.72 | | 0.48 |
| Education | -0.00007 | 0.00006 | -1.11 | 0.27 | | -0.00002 | 0.00001 | -1.32 | 0.19 | | -0.00002 | 0.00003 | -0.65 | 0.52 | | -0.00002 | 0.00003 | -0.64 | | 0.52 | | | -0.00002 | | 0.00003 | | -0.65 | | 0.52 |
| MMSE | 0.00003 | 0.00004 | 0.86 | 0.4 | | 0.00001 | 0.00001 | 1.12 | 0.27 | | 0 | 0.00002 | 0.05 | 0.96 | | 0.00002 | 0.00002 | 1.10 | | 0.28 | | | 0 | | 0.00002 | | 0.05 | | 0.96 |
| ZBI Total | 0 | 0.00001 | 0.22 | 0.82 | | 0 | 0 | -0.35 | 0.73 | | -0.00001 | 0 | -1.61 | 0.12 | | -0.00002 | 0.00001 | -2.80 | | 0.008 | | | -0.00001 | | 0 | | -1.61 | | 0.12 |
| Global Big 5 Discrepancy | 0.00016 | 0.00007 | 2.30 | 0.06 | | 0.00001 | 0.00002 | 0.37 | 0.71 | | -0.00006 | 0.00004 | -1.61 | 0.11 | | -0.00013 | 0.00004 | -3.07 | | **0.004** | | | -0.00026 | | 0.00004 | | -2.61 | | **0.022** |

**Table 5.** Effect of Big 5 Global Discrepancy on Right Hemisphere Cortical Volumes

|  | **Left Somatomotor** | | | |  | **Left Postcentral** | | | |  | **Left Frontal Eye Fields** | | | |  | **Left Inferior Frontal** | | |  | |
| --- | --- | --- | --- | --- | --- | --- | --- | --- | --- | --- | --- | --- | --- | --- | --- | --- | --- | --- | --- | --- |
|  | R2 | 0.12 |  |  | | R2 | 0.02 |  |  | | R2 | 0.081 |  |  | | R2 | 0.375 |  | | |
|  | F | 1.79 |  |  | | F | 0.796 |  |  | | F | 0.244 |  |  | | F | 6.01 |  | | |
|  | DF | 5, 52 |  |  | | DF | 5, 52 |  |  | | DF | 5, 52 |  |  | | DF | 5, 52 |  | | |
|  | p | 0.133 |  |  | | p | 0.513 |  |  | | p | 0.94 |  |  | | p | **0.00065** |  | | |
|  | β | SE | t | p | | β | SE | t | p | | β | SE | t | p | | β | SE | t | | p |
| Intercept | 0.00297 | 0.00126 | 2.36 | 0.02 | | 0.0017 | 0.00085 | 2.00 | 0.05 | | 0.00275 | 0.00197 | 1.40 | 0.17 | | 0.0013 | 0.00055 | 2.35 | | 0.023 |
| Age | 0.00002 | 0.00004 | 0.51 | 0.61 | | -0.00001 | 0.00001 | -0.79 | 0.44 | | 0 | 0.00002 | 0.02 | 0.98 | | 0.00001 | 0 | 1.39 | | 0.17 |
| Education | 0.00001 | 0.00002 | 0.55 | 0.59 | | -0.00003 | 0.00002 | -1.10 | 0.28 | | 0 | 0.00005 | 0.08 | 0.93 | | -0.00001 | 0.00002 | -0.73 | | 0.47 |
| MMSE | 0 | 0.00001 | -0.19 | 0.85 | | 0.00001 | 0.00002 | 0.78 | 0.44 | | 0.00003 | 0.00003 | 0.93 | 0.36 | | 0.00002 | 0.00001 | 1.62 | | 0.11 |
| ZBI Total | 0 | 0.00001 | -0.06 | 0.95 | | 0 | 0 | -0.68 | 0.5 | | 0 | 0.00001 | -0.30 | 0.76 | | -0.00001 | 0 | -2.26 | | 0.029 |
| Global Big 5 Discrepancy | -0.00001 | 0.00003 | -0.17 | 0.87 | | 0.00004 | 0.00003 | 1.40 | 0.17 | | -0.00001 | 0.00006 | -0.14 | 0.89 | | -0.00006 | 0.00002 | -3.23 | | **0.002** |
